# Effectiveness of COVID-19 Vaccines: A Vaccinated-Only Approach

**DOI:** 10.1101/2024.03.23.24304769

**Authors:** Ivo M. Foppa

**Affiliations:** Hessisches Landesamt für Gesundheit und Pflege, Außenstelle Dillenburg, Wolframstraße 33, 35683 Dillenburg, Hessen,Germany

**Keywords:** COVID-19, Vaccine effectiveness, Relative vaccine effectiveness, Quasi-exchangeability

## Abstract

We used a modified screening method that ensures quasi-exchangeability of comparison groups to estimate COVID-19 vaccine effectiveness in people resident in the Federal State of Hessen, Germany. COVID-19 vaccination history of vaccinated subjects with reported symptomatic SARS-CoV-2 infection was used to determine vaccination status. Subjects with their first COVID-19 vaccination within 7 days before the imputed date of infection were considered unvaccinated. Vaccination is assumed not to have a relevant effect on outcome risk for the first seven days and to be fully developed after between 14 and 21 days. The immunization profile of the source population was estimated from the number of subjects vaccinated by dose, date and age group as recorded in the Hessian COVID-19 vaccination registry. Effect estimates were obtained using logistic regression, fitted by a Bayesian approach. The first dose of COVID-19 vaccines had a measurable effect during the predominance of the Alpha and Delta variants of SARS-CoV-2, but a smaller effect during Omicron predominance. Only during Alpha and Delta predominance did the second dose provide an added benefit. During Omicron predominance, the third dose provided additional protection, but that effect was smaller than for the Delta period. Comparison of our estimates with estimates using a conventional, not quasi-exchangeable, approach revealed substantial differences in some cases, without any recognizable pattern.

**PACS:** 0000, 1111

**MSC:** 0000, 1111

**Graphical Abstract:** 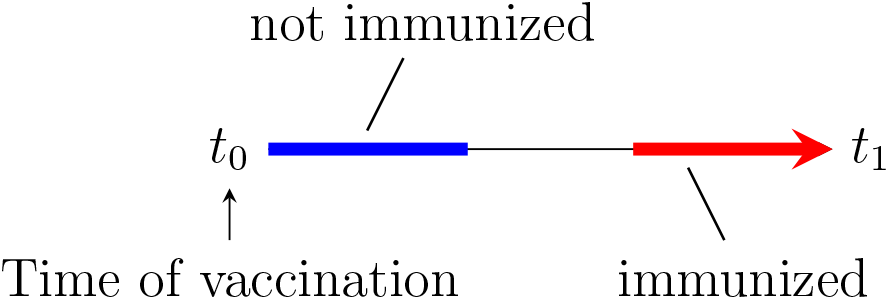

**Highlights:**

- We propose a vaccine effectiveness (VE) study design for COVID-19 that is based on a modification of the screening method. The modification ensures quasi-exchangeable: Only vaccinated subjects are considered, COVID-19 cases on the one hand and subjects registered in the COVID-19 vaccination registry. Risk comparisons are calculated for individuals who received their *n*-th vaccination one to two weeks apart, assuming no relevant vaccine effect within a week.
- The first dose of COVID-19 vaccines had a measurable effect during the predominance of the Alpha and Delta variants of SARS-CoV-2, but a smaller effect during Omicron predominance. Only during Alpha and Delta predominance did the second dose provide an added benefit. During Omicron predominance, the third dose provided additional protection, but that effect was smaller than for the Delta period.
- Comparison with a conventional approach revealed substantial differences in some cases, that did not follow a clear pattern.

## 1. Introduction

Presenting the largest global public health crisis in over 100 years, the severe acute respiratory syndrome corona virus 2 (SARS-CoV-2) pandemic of 2020 through 2023 triggered an effort to develop and approve vaccines at an unprecedented pace [1]. These vaccines included mRNA and vector-based formulations. Randomized clinical trials indicated excellent efficacy for these vaccines, up to over 90% [2]. Several of these vaccines are now approved for use in many countries by national and supra-national regulatory bodies [3]. Massive vaccination campaigns in the context of a soaring pandemic provide a unique opportunity to study post-licensure effectiveness of these vaccines. This is even more important, as randomized clinical trials for these vaccines are no longer ethically feasible in the groups for whom effectiveness data is available. The most widely used study design for assessing the effectiveness of vaccines, in particular vaccines against influenza, is the test-negative design (TND) [4]. In the case of influenza, this modified case-control design offers clear advantages over other observational designs, most importantly by reducing a health care-seeking bias. In fact, the World Health Organization recommends that design as the least biased observational design for studying the effectiveness of COVID-19 vaccines and also “[…] the most feasible approach in most settings.” [5, p. 4015] and numerous recent studies have used a TND to estimate COVID-19 vaccine effectiveness (VE) (see, e.g., [6, 7, 8, 9, 10]). However, there are concerns regarding the suitability of the TND to study COVID-19 VE, which we will present elsewhere. These concerns revolve around the very limited circulation of respiratory viruses other than SARS-CoV-2 during much of 2020/21 and the more specific clinical presentation of COVID-19 compared to influenza.

Uphoff et al. [11] proposed a “case-series approach” to study the effectiveness of an AS03-adjuvanted vaccine against influenza A(H1N1)pdm2009. Here, we present a study that is based on the principle proposed by Uphoff and colleagues, but uses a screening method approach. By only using vaccinated cases, biases that plague other observational designs are mitigated. This is true because of a reasonable assumption of exchangeability [12] regarding subjects of different exposure status. Thus, we propose our modification of the screening method to represent a *quasi-randomized* approach.

## 2. Methods

### 2.1. Study design

Our study used a modification of the screening method [13]. Briefly, this design, which is conceptually related to a case-control design, also uses cases, but exposure prevalence in the source population is estimated using “external” data, e.g. from a population survey. Our study differed in important ways from standard implementations of this design:

1. Only vaccinated subjects were used.
2. Reported cases of COVID-19, all vaccinated, were classified as “not immunized”, when the first vaccination against SARS-CoV-2 was up to 7 days before the imputed date of infection and as “immunized” by the first dose when vaccination had been 15 to 21 days before.
3. Similar determinations where made regarding the *n*-th dose of the vaccine: If the imputed date of infection was Up to 7 days after the *n*-th vaccination, cases were considered effectively immunized with *n* – 1 doses of the vaccine; with a date of infection between 15 to 21 days after vaccination with the *n*-th dose they were considered immunized with *n* doses.
4. The assumed source population was constructed from the number of subjects having received a COVID-19 vaccine in Hessen, after classification into exposure categories equivalent to the cases.

Vaccination effects were estimated from the comparison between “immunized” and “not immunized” after the *n*-th dose of the vaccine, for *n* ∈ 1, …, 4. Thus, for the first dose, VE_1_ is estimated while, for doses 2 through 4, relative VE (rVE_2_ through rVE_4_) are estimated. As stated above, comparison is always between those who had received their *n*-th dose of the vaccine up to 7 days before the imputed date of infection (not yet immunized by latest vaccine dose) and those who had received that dose between 14 and 21 days before (immunized by latest vaccine dose).

### 2.2. Data

The cut-off date used for this study was 3-24-2022. Data from two sources were used for this study:

#### 2.2.1. Outcome data

Vaccinated cases of SARS-CoV-2 infection in the German federal State of Hessen were obtained from the the German mandatory infectious disease reporting system. Only cases for whom a date of illness onset was recorded, who met the German reference definition (positive PCR test [14]), who were vaccinated with a vaccine licensed in Germany [15], with a plausible vaccination date (the vaccination campaign started on December 27, 2020) at or before the imputed date of infection *d*^*^ (main analysis) or date of symptom onset *d* (secondary analysis), who had a reporting date after the imputed date of infection and who were at least 12 years old at that time were included in the analysis. The imputed date of infection for subject *i* was calculated As 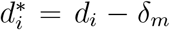, where *δ*_*m*_ was the incubation assumed for the given time period. For the time period from December 27, 2020 through July 4, 2021, when the Alpha variant (B.1.1.7) of SARS-CoV-2 predominated in Hessen, we assumed an incubation period of four days [16]. For the following time period that lasted until January 2, 2022, when the Delta (B.1.617.2) predominated and after that, when the Omicron variant ((B.1.1.529) predominated, an incubation period of three days was assumed [17, 18]. For each case *i*, the number *d*_*i*_ of COVID-19 vaccinations received and the date *t*_*i*_ of the last vaccination was given. Accordingly, cases were classified according to their immunization status:

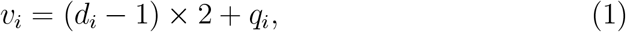

where *q*_*i*_ is an indicator variable for time since vaccination, i.e.

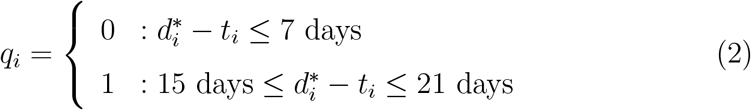

Thus, e.g., those with an imputed date of infection 0 to 7 days after receiving the second dose vaccination would be attributed an immunization category of *v*_*i*_ = 2 and those between 15 and 21 days after there second dose would be considered *v*_*i*_ = 3. Cases with a vaccination date 8 to 14 days before their imputed date of infection were discarded.

Cases with immunization status *v*_*i*_ ∈ 0, 2, 4, 6 were considered not effectively immunized by their latest dose of vaccination (“unexposed”) and *v*_*i*_ ∈ 1, 3, 5, 7 were considered immunized by their first, second, third or fourth dose, respectively (“exposed”).

#### 2.2.2. Vaccination cohort

The daily number of people by age group (12 to 17, 18 to 59 and 60+ years old) receiving their first, second, third or fourth dose of a COVID-19 vaccine approved in Germany at the time [15], were obtained from daily vaccination numbers for Hessen. These numbers were compiled daily by the Robert Koch Institute [19] and are known to under-represent the administered vaccination doses by no more than 5 percentage points [20]. From these numbers we computed daily numbers of people in the immunization categories that were defined for the cases. For example, for date *t*, we calculated the numbers of subjects of age category *a*, not immunized at all, *n*_0,*a*_(*t*) as

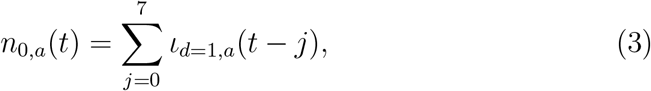

where *ι*_*d*=1,*a*_(*t*) is the numbers of first doses of the vaccine administered on date *t* to subjects of age group *a*. Similarly, the number of subjects effectively boostered for the second time was calculated as

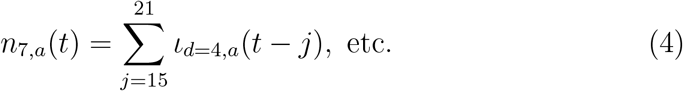

#### 2.2.3. Periods of predominance of SARS-CoV-2 variants

We defined periods of SARS-CoV-2 variant predominance by the start of the first week during which the majority of samples characterized by either whole-genome sequencing or melting point analysis were classified as a particular SARS-CoV-2 variant. In Germany, including Hessen, the COVID-19 vaccination campaign started in 2020-12-27. At the time, the SARS-CoV-2 Alpha variant (Pangolin B.1.1.7) predominated. The Alpha predominance ended on 2021-07-04 and was succeeded by the Delta variant (B.1.617.2), which was followed by Omicron, starting on 2022-01-02.

### 2.3. Statistical analysis

#### 2.3.1 Logistic model

We fit logistic regression models of the form:

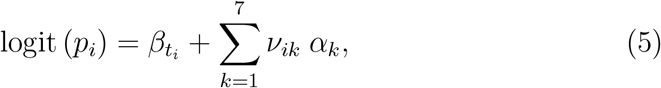

where *p*_*i*_ is the probability that study subject *i* with index date *d*_*i*_ and vaccination status *v*_*i*_ is a case, *c*_*i*_ = 1, and 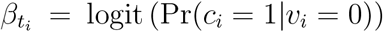 is the random intercept. Vaccination status is encoded by the dummy variables *ν*_*ik*_, *k* ∈ 1, …, 7 and vaccination effects are captured by the coefficients *α*_*k*_, *k* ∈ 1, …, 7. The vaccination status dummy variables were coded in the following manner:

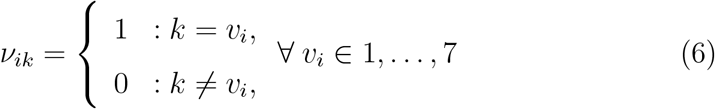

and

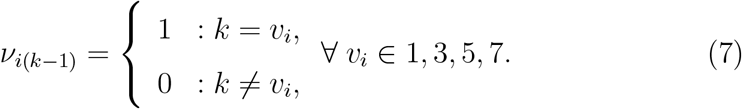

Thus, the likelihood contribution of a case *i* of immunization status *v*_*i*_ = 1, e.g., i.e. between 15 and 21 days after receiving the first dose, is

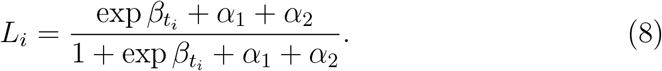

The coefficient estimate of *α*_2_ can be used to calculate the estimate of VE_1_,

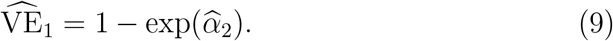

Equivalently, the estimates of the coefficients *α*_3_, *α*_5_ and *α*_7_ can be used to calculate estimates for vaccination benefits, but here these estimates represent rVE_2_, rVE_3_ and rVE_4_, respectively:

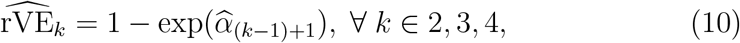

where rVE_*k*_ is short for rVE_*k* vs.*k*−1_, the relative effectiveness of dose k compared to dose *k* − 1 at the time of receiving the *k*th dose, as measured in dose *k* recipients within 7 days. E.g., rVE_2_ is the relative VE of the second dose, after the time deemed necessary to develop its optimal effect (between 15 and 21 days), compared to the first dose at time of receiving the second dose (≤ 7 days).

The coefficients *α*_2_, *α*_4_ and *α*_6_, however, cannot be meaningfully interpreted in terms of VE. For the comparison presented in section 3.2 we will refer to this approach as “quasi-exchangeable analysis”.

#### 2.3.2. Model fitting

Because of sparse data we used Bayesian methods, specifically the R2jags package [21] in R [22] under Rstudio [23] to fit these models. Age group– dose–period strata for which no doses were administered in Hessen or in which there were fewer than ten cases were discarded. All model parameters were assigned flat Normal priors N(0, 1e−6). After a run-in period of 10,000 iterations we took 10,000 samples, using every fifth for inference. Analyses were conducted separately for the three age groups and for the periods defined by the predominance of different variants of SARS-CoV-2.

### 2.4. Conventional analysis

To compare results from our analysis with results obtained by a conventional approach, which is based on the comparison of outcomes in vaccinated vs. unvaccinated subjects. To that end, we defined daily cases similarly as described above, but with the following differences: Cases were classified as vaccinated if they had received a vaccine at least 14 days before their imputed date of infection. If they had received their *k*th dose of a COVID-19 vaccine within that 14-day window, they were classified as vaccinated by *k* − 1 doses; e.g. they would be considered non-vaccinated after their first dose. The number of vaccinated controls at a given date, by age group, was the cumulative number of persons with a 14-day lag. The number of persons vaccinated once was then calculated by subtracting the cumulative number of second dose recipients, again lagged, from the cumulative number of once vaccinated etc. For the middle age group this resulted negative values for the number of subjects vaccinated once during the last 30 days of the data (starting day 440 after beginning vaccination). We therefore only analyzed the first 430 days. The data was analyzed as described but using the following modified logistic regression model:

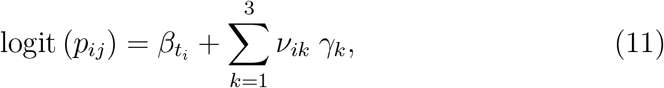

where

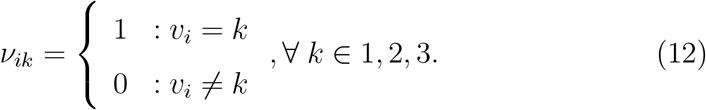

VE estimates for the first, second and third dose were calculated as VE_*k*_ = 1 − exp(*γ*_*k*_).

## 3. Results

The numbers of vaccine doses administered in Hessen to different agegroup-dose-period strata varied widely (Table 1). While, e.g., during the predominance of the SARS-CoV-2 Alpha variant the majority of doses were administered to subjects not yet vaccinated, during the Omicron period the most doses, by far, were booster doses. Booster doses were not yet available during the Alpha period. During the Omicron period, first and second doses were administered only rarely to people 60 or older in Hessen because most of them were already fully vaccinated.

**Table 1:** Numbers of cases and number of doses administered in Hessen, Germany, by age group, most recent dose of a COVID-19 vaccine and predominant SARS-CoV-2 variant. The cases are restricted to those with their latest dose of the vaccine received less than three or between 15 and 21 days before imputed date of infection.

| Age Group | Dose | Alpha |  |  | Delta |  |  | Omikron |  |
| --- | --- | --- | --- | --- | --- | --- | --- | --- | --- |
|  |  | Cases (total) | # Doses |  | Cases (total) | # Doses |  | Cases (total) | # Doses |
| 12 to 17 | 1 | 6 (10) | 46,635 |  | 150 (369) | 194,665 |  | 86 (643) | 22,963 |
|  | 2 | 0 (3) | 12,100 |  | 77 (762) | 190,735 |  | 93 (3,401) | 36,121 |
|  | 3 | 0 (0) | 0 |  | 4 (7) | 14,545 |  | 195 (2,861) | 92,860 |
|  | 4 | 0 (0) | 0 |  | 0 (0) | 0 |  | 1 (25) | 1,899 |
| 18 to 59 | 1 | 918 (1,869) | 1,954,323 |  | 945 (5,390) | 801,340 |  | 255 (6,441) | 75,467 |
|  | 2 | 78 (281) | 1,279,002 |  | 587 (21,342) | 1,395,556 |  | 460 (19,317) | 173,639 |
|  | 3 | 0 (1) | 42 |  | 983 (1,782) | 1,277,021 |  | 1,832 (58,805) | 945,093 |
|  | 4 | 0 (0) | 0 |  | 0 (25) | 58 |  | 120 (1,308) | 105,503 |
| 60+ | 1 | 518 (989) | 1,444,083 |  | 60 (461) | 120,096 |  | 66 (1,658) | 10,510 |
|  | 2 | 78 (235) | 1,075,086 |  | 54 (5,484) | 465,637 |  | 45 (2,054) | 25,009 |
|  | 3 | 0 (1) | 12 |  | 292 (663) | 1,152,134 |  | 223 (15,052) | 320,758 |
|  | 4 | 0 (0) | 0 |  | 0 (2) | 22 |  | 275 (2,675) | 418,930 |

\* Most recent dose of a COVID-19 vaccine.
† The number of doses of a COVID-19 vaccine administered as first, second or third dose, respectively, in that particular age group and period.
‡ Vaccine not yet recommended by the German Standing Committee on Immunization (Ständige Impfkommission, STIKO).

**Table 2:** Estimates of VE_1_ and estimates of VE_2_, VE_3_ and VE_4_, calculated using (9) and (10), for the three age groups and periods, respectively.

| Age Group | VE | Predominant SARS-CoV-2 Variant |  |  |
| --- | --- | --- | --- | --- |
|  |  | Alpha | Delta | Omikron |
| 12 to 17 | $VE_1$ | . | 60.5 (46.8,72.8) | -84.8 (-171.9,-18) |
| | $rVE_2$ | . | 68.1 (51.5,81.6) | -51 (-118.2,1.2) |
| | $rVE_3$ | . | . | 28.6 (8,47.4) |
| | $rVE_4$ | . | . | . |
| 18 to 59 | $VE_1$ | 53.7 (47.2,60.4) | 38.2 (30.9,46) | -14.8 (-41.7,11.2) |
| | $rVE_2$ | 83.3 (70.4,92.7) | 61.6 (55.6,67.9) | -36.5 (-59.4,-13) |
| | $rVE_3$ | . | 59.1 (53.6,64.9) | -43.3 (-56.6,-29.4) |
| | $rVE_4$ | . | . | 9.9 (-22.2,37.5) |
| 60+ | $VE_1$ | 40.1 (29.9,50.3) | 48.8 (20.2,70.3) | 58.6 (34.7,77) |
| | $rVE_2$ | 88.2 (78.2,95.2) | 75.9 (59.5,88.1) | -77.7 (-196.1,4.4) |
| | $rVE_3$ | . | 70.2 (62,77.8) | -22.7 (-56.3,8.1) |
| | $rVE_4$ | . | . | 18.2 (-0.3,35.8) |

\* Vaccine not yet recommended by the German Standing Committee on Immunization (Ständige Impfkommision, STIKO).
† 95% credible interval.

### 3.1. Estimates of VE_1_, rVE_2_, rVE_3_ and rVE_4_

During the predominance of the Alpha SARS-CoV-2 variant, all estimates of VE_1_ and rVE_2_ (only age groups 18+) were statistically signficantly positive, indicating (relative) effectiveness (Table **??**). The same was true for the period of Delta SARS-CoV-2 variant predominance, when also VE_1_ and rVE_2_ estimates were available for the youngest age group as well as rVE_3_ estimates for the older age groups. During the Delta period, most estimates were lower compared to the Alpha period, although only some differences were statistically significant. During the Omicron period, most estimates were lower than for the two other periods and, in fact, lacked statistical evidence of any adventageous effect. The two exceptions were the rVE_3_ estimate for the lowest age group and the VE_1_ estimate for the oldest age group.

### 3.2. Methods comparison

Our approach allowed for the estimation of VE_1_, the effectiveness of the first dose, but only for the estimation of the relative effectiveness of the second and following doses, with respect to the previous dose, rVE_2_ and rVE_3_, respectively. Even though these quantities are of interest themselves, as they capture the added benefit of receiving a second and third dose of the vaccine, the overall effect of a vaccination regime may greater interest. We therefore compared only VE_1_ estimates (Figure 1). For the youngest age group, estimates could not be obtained during the Alpha period. For the other age groups, VE_1_ from the conventional analysis was in the mid-nineties, from the quasi-exchangeable analysis only slightly above 60%. While the period-specific estimates of VE_1_ clearly declined from the earlier periods to the Omicron period, although not always statistically significant, such decline more subtle for the conventional analysis in general, and for the 60+ almost entirely absent. The conventional estimates of VE_1_ were always drastically and statistically significantly (*α* = 0.05) higher than the quasi-exchangeable ones.

**Figure 1:**
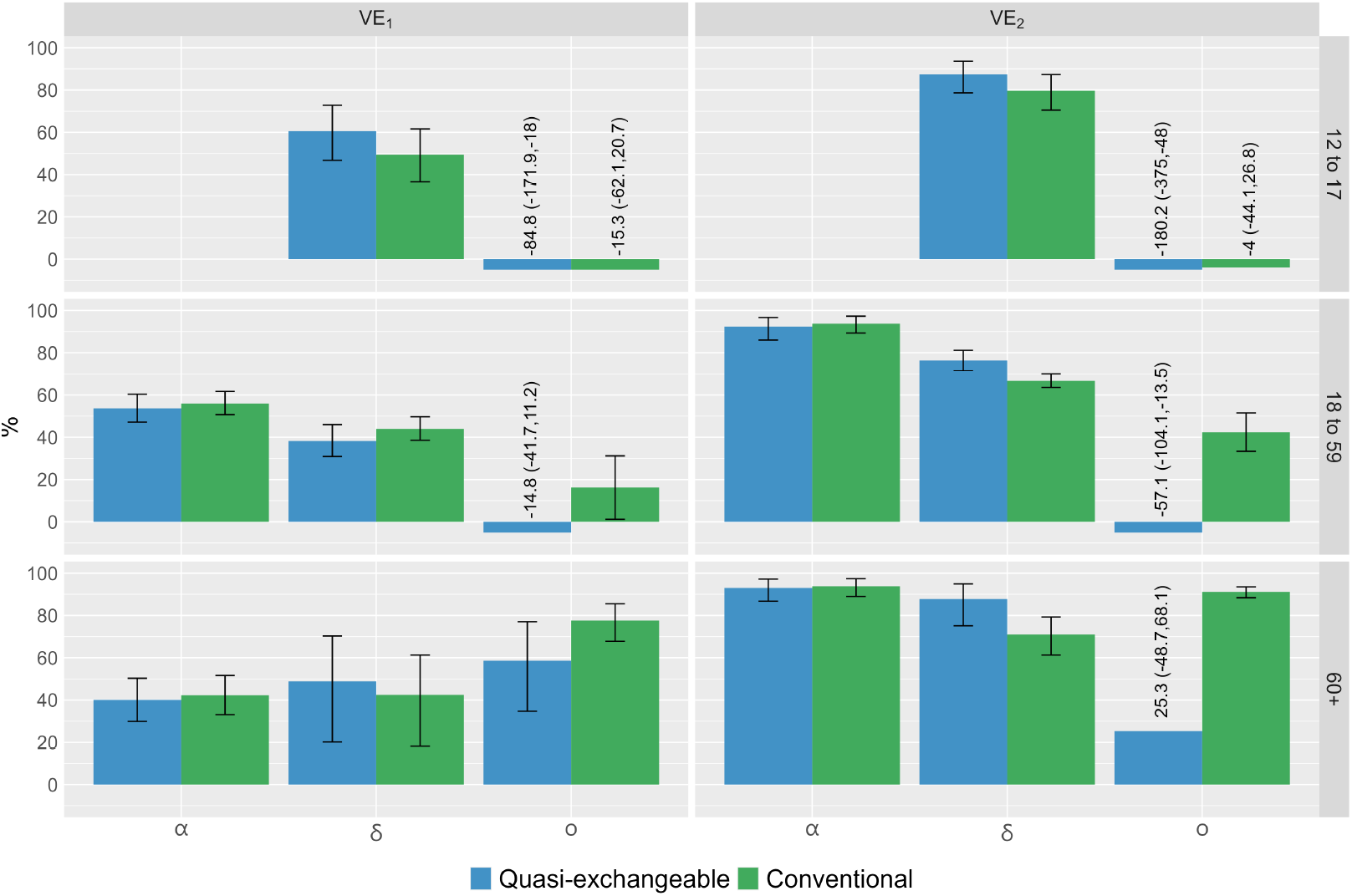
Estimates of VE_1_ und VE_2_ during the three periods of SARS-CoV-2 variant predominance, by age group and method (quasi-exchangeable vs. conventional analysis– see text for details). Error bars, representing 95% credible intervals, that would extend beyond the limits of the y-axis are replaced with numbers.

## 4. Discussion

One major issue of observational studies of outcomes of vaccination is confounding by indication [24] that results from treatment assignment based on prognostic factors. Furthermore, the receipt of a vaccine against COVID-19 is based on a voluntary decision, which may be informed by factors likely associated with risk/risk avoidance behaviors [25, 26], adding to potential confounding of observational VE estimates. To eliminate such sources of bias, we restricted our study to only vaccinated subjects.

We consider this design to be quasi-randomized, because exposure can reasonably be assumed to be independent of unmeasured confounders as all “exposed” received the same treatment as the “unexposed”: a first, second or third dose of a SARS-CoV-2 vaccine. However, the assumption of strict exchangeability [12] would be hard to justify as exposed and unexposed subjects differ with respect to vaccination date: COVID-19 vaccine hesitancy is not random [see, e.g., 27] and therefore, people receiving their first dose of a COVID-19 vaccine now are, on average, different from those who rushed to get their first shot at a different stage of the SARS-CoV-2 pandemic. Such differences could include behavioral differences that inform risk of infection. However, as the comparison groups differ with respect to the date of receipt, *t*, of their *k*th dose of COVID-19 vaccine by only up to 21 days it seems safe to assume that systematic differences between those groups are minimal. We will therefore consider these comparator groups as *quasi-exchangable* and justify that in the Appendix. That condition cannot necessarily be claimed between other groups that differ with respect to immunization status.

According to our approach, levels of “exposure” are defined by time since the receipt of a given dose of a COVID-19 vaccine. In order for effect estimates to be valid, exchangeability criteria need not to be met for vaccinated and unvaccinated subjects, which, in an observational setting would be unlikely to hold. Rather, the criterion needs to be met by groups differing with respect to the time of receipt of a given dose of the vaccine, our levels of exposure. While it is quite obvious that subjects who received their first dose of a COVID-19 vaccine right after licensing differ, on average, in their relevant behavior from subjects who receive their first dose now under the threat of vaccination mandates, exchangeability could not reasonably be claimed for these groups. It is therefore evident, that time of vaccination may confound VE estimates that are based on exposure categories defined by time since vaccination. Yet, in our case the time difference that separates “exposed” and “unexposed” subjects ranges from 12 to 21 days. We have not data to validate the assumption that such difference is irrelevant to prognostic differences among comparison groups. However, we claim that such differences will be relatively small, in particular when compared to differences between vaccinated and unvaccinated subjects, as it is done in the most widely used design of observational VE, the test-negative design [4]. While not claiming true exchangeability of comparison groups in our design we use the term *quasi-exchangeability*.

The question regarding the biological correlate of this result can, obviously, not be answered with our data. It should be pointed out, though, that the first dose of a mRNA COVID-19 vaccine has previously found to be very effective in adolescents[28]

## Data Availability

All data used in the present study are available in aggregated form upon reasonable request to the authors

https://www.rki.de/DE/Content/InfAZ/N/Neuartiges_Coronavirus/Daten/Impfquoten-Tab.html

## Appendix A Calculating VE_2_ from rVE_2_ and VE_1_

Let

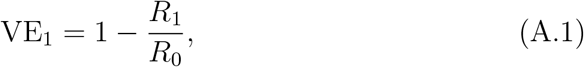

where *R*_1_ is the “risk” of those vaccinated once with COVID-19 vaccine and *R*_0_ is the “risk” in the unvaccinated. The term “risk” is used here to represent any measure of (disease) occurrence, such as attack rate, cumulative incidence or hazard rate. Similarly, let

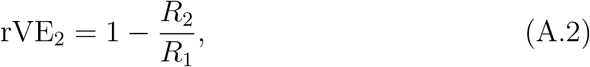

where *R*_2_ is the risk in the fully vaccinated, after attaining optimal immunity (after *>* 14 days) and *R*_1_ is the risk before the second dose is assumed to have any effect on risk of infection (≤ 3 days after vaccination).

We can rearrange (A.1) and (A.2) to obtain expressions for *R*_0_,

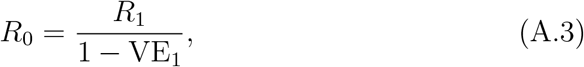

and for *R*_2_

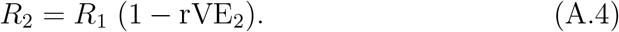

Substituting these expressions into (A.5) we obtain an expression for VE_2_ in terms of VE_1_ and rVE_2_

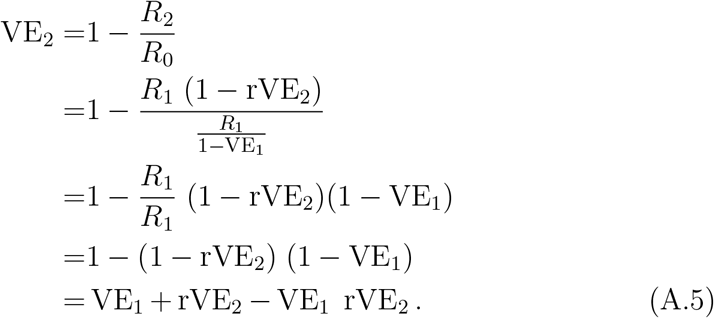

